# A study of sex-specific genetic effects underlying risk of orofacial clefts also highlights the potential impact of sequencing errors due to short read mis-mapping

**DOI:** 10.64898/2026.07.07.26357463

**Authors:** Kanika Kanchan, Zeynep Erdogan-Yildirim, Seth Berke, Nandita Mukhopadhyay, Debashree Ray, Claire L Simpson, Jacqueline A Bidinger, Sarah W Curtis, Azeez Butali, Holger Schwender, Alan F Scott, Joan E Bailey-Wilson, Terri H Beaty, Elizabeth Leslie-Clarkson, Mary L Marazita, Ingo Ruczinski

**Author notes:** To whom correspondence should be addressed: **Mary L Marazita**, Center for Craniofacial and Dental Genetics, Department of Oral and Craniofacial Sciences, School of Dental Medicine, University of Pittsburgh, Bridgeside Point, Suite 400, 100 Technology Drive, Pittsburgh, PA 15219. **Ingo Ruczinski**, Department of Biostatistics, Johns Hopkins Bloomberg School of Public Health, 615 North Wolfe Street, Baltimore MD 21205.

## Abstract

Orofacial clefts (OFCs), including cleft lip (CL), cleft palate (CP), and cleft lip with cleft palate (CLP), are among the most common craniofacial malformations in humans, with a birth prevalence of approximately 1 in 1,000 live births globally. Non-syndromic forms of OFC are predominantly genetic, with significant variability in prevalence across populations. Understanding the genetic underpinnings of OFCs remains a key public health priority, given the substantial medical and societal burden of these conditions. Recent genome-wide association studies (GWAS) have implicated numerous genetic loci, but challenges remain due to genetic heterogeneity and complex gene-environment interactions. This study aimed to identify sex-specific genetic risk factors for cleft lip with or without cleft palate (CL/P) through a meta-analysis of whole genome sequencing (WGS) data from 1,922 case-parent trios across eight diverse cohorts. Our approach revealed four SNPs in three distinct regions that showed genome-wide significant sex-specific effects. However, despite each of these SNPs passing standard quality control filters, follow-up analyses showed that these signals most likely were technical artifacts caused by sequencing errors, in particular mis-mapped reads due to sequence similarities with the sex chromosomes. These findings highlight the necessity for careful scrutiny when studying differences between the sexes in genetic association studies.

## Introduction

Orofacial clefts (OFCs) represent the most common group of craniofacial malformations in humans affecting approximately one per 1,000 live births worldwide ^1^. OFCs include cleft lip (CL), cleft palate (CP) and cleft lip with cleft palate (CLP), which can occur as isolated malformations, with another malformation or as part of a recognized malformation syndrome (often Mendelian with some incomplete penetrance). OFCs are commonly categorized into two anatomically and embryologically distinct entities based on embryologic and epidemiologic patterns: cleft lip with or without cleft palate (CL/P) and cleft palate alone (CP) ^2^. Among all infants born with an OFC, 70 percent of CL/P cases and 50 percent of CP cases occur as isolated, non-syndromic malformations ^3^. Non-syndromic CL/P occurs more frequently in males than females (male to female ratio approximately 2:1) whereas non-syndromic CP occurs more often in females (male to female ratio approximately 1:1.14) ^4,5^. Substantial variation in birth prevalence rates of non-syndromic CL/P has been reported across populations: Asian populations have higher birth prevalence rates compared to European populations ^6^ and African populations have the lowest birth prevalence rates ^5^. Non-syndromic CP also shows variability in birth prevalence rates across populations, although issues of misclassification of CP may influence reported prevalence rates in low to middle income countries ^7,8^. Collectively OFCs represent a major public health burden for patients and their families, the health care system and society^9^. Due to the high overall birth prevalence rate and the large financial, medical and emotional burden of treatment required by children with an OFC, understanding their etiology remains an important public health goal.

Risk to OFC shows strong evidence of genetic control with population-based twin and family studies suggesting heritability up to 90% based on twin registry data from Europe ^10^. Genetic studies (both twin and family studies) date back more than two hundred years ^11^. Despite this long history of scientific research into the genetic control of OFC including both linkage ^12^ and association studies ^13,14^, it remains difficult to clearly identify underlying causal genes. This may reflect the genetic heterogeneity influencing risk to OFC, where a number of different genes (with both rare and common variants) control risk^13,15^. Recent genome-wide linkage and association studies have clearly shown that multiple genes play a role in the etiology of OFCs, but with substantial heterogeneity among families and across populations. To date, approximately 50 different genes have been identified as significant in genome-wide studies of OFCs ^13^, with about two dozen having substantial replication and/or functional studies. Causal genes for Mendelian malformation syndromes including OFCs as a key phenotype should also be used to identify candidate genes (see Table 1 in Seto-Salvia et al. ^16^ for a list) although very few common polymorphic variants within these apparently causal syndromic genes show any evidence of association with non-syndromic OFC in population-based studies. This high level of genetic heterogeneity for OFCs occurs on top of recognized environmental risk factors (e.g. maternal smoking during pregnancy, nutrition, etc.) which raises the possibility that gene-environment interactions may also be important. Thus, OFCs are clearly complex (i.e. influenced by both genetic and non-genetic factors) and heterogeneous (with multiple genes involved).

**Table 1:**
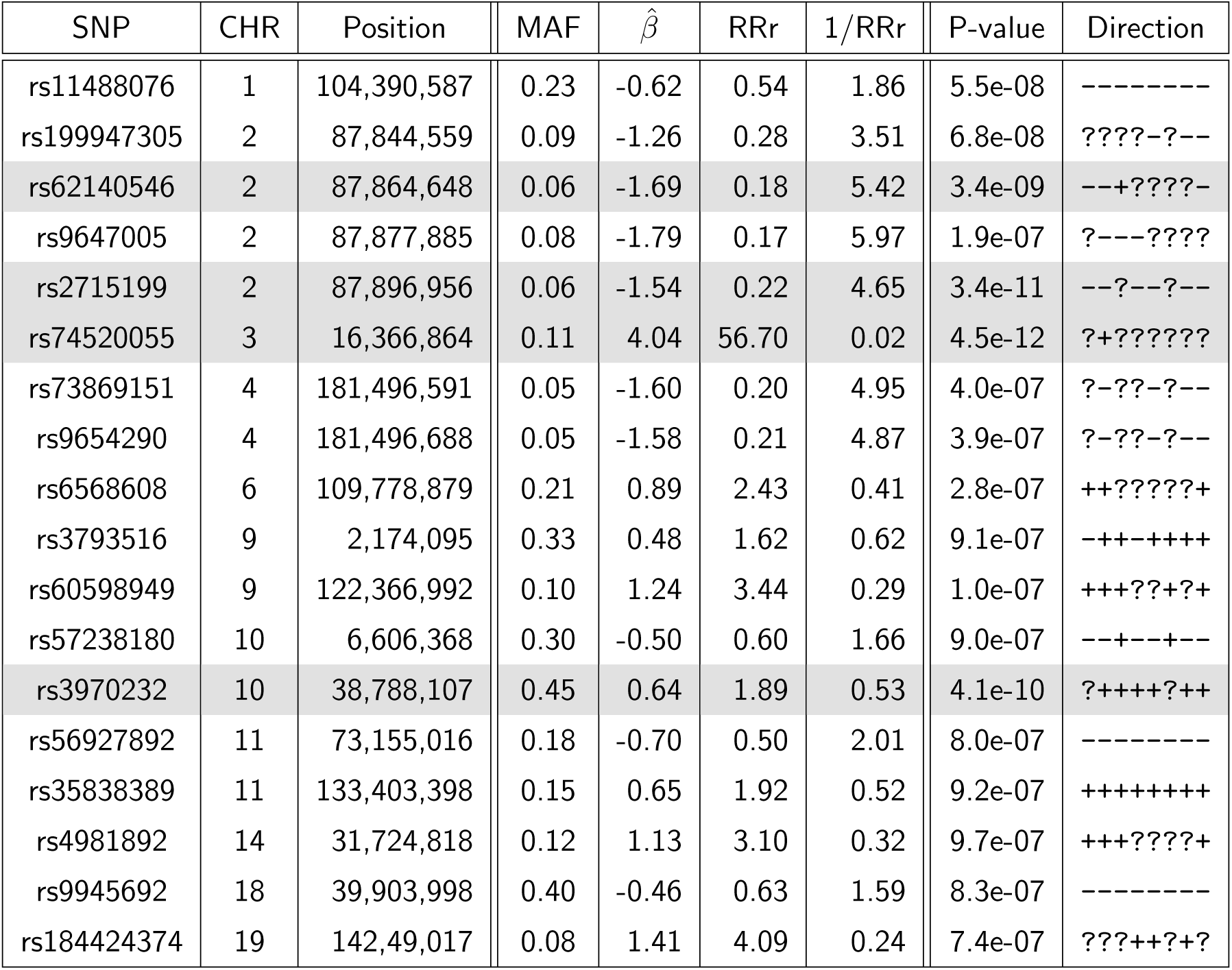
Variants with a p-value less than 1 × 10^−6^ in the meta-analysis, testing the 1 degree of freedom SNP *×* Sex interaction in the conditional logistic gTDT model. The variants passing the genome-wide significance threshold of 1 × 10^−8^ are highlighted with a grey background. The columns for each SNP show the variant rsID (SNP), the chromosome (CHR) and chromosome location (Position), the meta-analysis minor allele frequency across the studies (MAF), the estimate of the interaction parameter from the gTDT (*β*^^^), the relative risk ration (RRr) comparing male carriers of the variant allele to female carriers and its reciprocal (1/RRr) comparing female carriers to male carriers, the p-value and the direction of the effect. A ‘+’ indicates a positive slope estimate in the respective study, a ‘-’ indicates a negative slope estimate and the ‘?’ indicates that the variant was not included in the meta-analysis (being monomorphic in the study, failing QC or having a minor allele frequency below 1%). These signs reflect the cohorts in the order as shown in Supplementary Tables 1 – 3.

The case-parent trio design tests a composite null hypothesis of no linkage or no association so marker transmission from a heterozygous parent to an affected child should always be the expected 50% under this null ^17^. A key advantage of this trio study design, which allows us to distinguish between transmitted and non-transmitted parental alleles, is its robustness to the inflated Type 1 errors due to population stratification possible in case-control studies^18^. In addition, the case-parent trio design makes it easy to incorporate tests of gene-environment interaction ^19^. In particular, genome-wide analyses of such gene-environment interactions can rapidly be carried out when the environmental variable is binary (e.g., maternal smoking yes/no) as closed form solutions exist for the estimates necessary for statistical inference ^20^. Thus, this approach can also be used to assess allelic heterogeneity (SNP*×*Sex interactions) by coding sex as a binary variable.

There are multiple hypotheses behind sexual dimorphism in cleft type and prevalence rates. For instance, reduced risk of CL/P in females could be due to: 1) a compensatory copy of chr X or a lack of chr Y in females; 2) differential liability thresholds of risk; 3) differential effects of autosomal variants due to hormonally mediated dimorphism; 4) effect measure modification by sex. A sex-stratified study of CLP in China found stronger association with markers in 1q32.2 in males compared to females, consistent with the higher prevalence of CLP in males ^21^. In separate case-control studies, our collaborators have previously examined if effects of risk alleles for OFCs are modified by sex and found suggestive evidence (p *<* 10^−5^) at 17 loci in a European ancestry cohort ^22^ and 93 loci in African ancestry cohorts ^23^. Most GWAS on OFCs however have either adjusted for sex as a main effect^24,25^ only or ignored sex altogether^26–31^ (including studies using a trio design e.g., ^19,32–35^) in a combined analysis of the sexes, likely due to limited sample size and power of stratified analysis to identify autosomal variants with sex-differential effects. Here, we comprehensively tested for sex differences in autosomal genetic risk effect sizes in a large number of multi-ethnic case-parent trios, conducting a meta-analysis on whole genome sequences from a total of eight cohorts.

## Materials and Methods

### Cohorts

Data from eight cohorts were analyzed in this manuscript (Supplementary Table 1). The data analyzed from cohorts 1, 2, 5, 7 and 8 are from participants drawn from the multi-ancestry Pittsburgh Orofacial Cleft study (POFC) including a variety of simplex and multiplex pedigrees of varying sizes and geographic origin. The data analyzed from cohort 4 are from the International Consortium to Identify Genes and Interactions Controlling Oral Cleft, part of the larger Gene Environment Association Studies (GENEVA) program. For the current study only complete case-parent trios were included. Detailed descriptions of sample collection are available in previous publications ^27,35–39^.

### Quality control procedures

Trios were sequenced at the Broad Institute with the resulting multi-sample variant call format (VCF) files for case-parent trios released by the GMKF Data Resource Center (DRC) at the Children’s Hospital of Philadelphia (all cohorts except 6) or sequenced and released by the Center for Inherited Disease Research (CIDR) at Johns Hopkins University (cohort 6). Sequencing details and data processing are given in previous publications ^35,38,40^. The same following QC pipeline was applied to all complete trios in the VCF data sets before being analyzed. We used VCFtools ^41^ to remove multi-allelic sites and indels, filter sites with labels other than ‘PASS’, to remove variants with mean depth values less than 10 or quality values less than 20, and to remove genotype calls with less than 10X read depth or quality less than 20. Binary PLINK^42^ files were created from the cleaned VCFs to integrate the pedigree information of the affected probands. Using PLINK functionality, we excluded sites with more than 20% missing genotypes, minor allele frequency (MAF) below 1%, and those significantly deviating from Hardy Weinberg Equilibrium with a p-value less than 10^−6^. Trios were exluded from the analysis if they had at least one sample with more than 2% missing genotypes, if they produced excessive Mendelian error rates or showed ambiguous relatedness based on pairwise IBD estimates. To derive these estimates, we first performed chromosome-wise linkage disequilibrium (LD) pruning in PLINK using variants with MAF larger than 5%. Pairs of variants in a window of 100 variants with R^2^ greater than 0.2 were noted, and variants were greedily pruned from the window until no such pairs remained. LD-pruned variants of all chromosomes were merged into a single file and IBD was estimated for all pairs of individuals in the dataset. Siblings among affected probands were identified through these IBD estimates and separate case-parent trios were encoded for subsequent analysis.

### Testing SNP *×* Sex associations

We used the genotypic transmission disequilibrium test ^43^ (gTDT) for the analysis of the case-parent trios. At each marker, one of four possible pairs of parental alleles is transmitted to the affected offspring, and the other three unobserved genotype realizations can be used as artificial controls (usually referred to as pseudo-controls). The resulting matching structure is accounted for using a conditional likelihood ^44^, which yields parameter estimates and estimated standard errors and enables the direct assessment of relative risk (RR) ^44,45^. In addition to the main (genotype) effect we also included a SNP *×* Sex interaction, which allows to test for differences in genetic effect sizes between the sexes. In standard implementations the parameters and their respective standard errors in this conditional logistic regression model are estimated numerically using an iterative procedure. However, we previously showed that Mendel’s laws impose a structure on the genotypes that allows us to derive exact closed-form solutions for the parameter estimates and estimated standard errors in this conditional logistic regression model, including the SNP *×* Sex interaction ^20^. This dramatically reduces computing time by avoiding iterative numeric optimization and makes genome-wide scans scalable for millions of markers. In this manuscript we used this procedure, implemented in the freely available Bioconductor package trio^46^.

### Meta-analyses

The SNP *×* Sex interaction summary statistics were meta-analyzed using METAL^47^. The software implements an inverse-variance weighted meta-analysis method, which combines effect size estimates from multiple cohorts while accounting for their standard errors, thus maximizing statistical power and accounting for sample size variability. By tracking reference and alternate alleles, the software ensures consistent aggregation of results across studies. For each cohort, only SNPs with MAF 1% or larger were included in the meta-analyses. Genomic control^48^ of test statistics and p-values was employed in METAL for each cohort and for the meta-analysis p-values.

### Post-analysis quality control

We used BLAT^49^ and the self-chain alignments ^50^ from the UCSC Genome Browser ^51,52^ to identify regions of similarity between the DNA sequence around our variants of interest and the hg38 reference genome, including the sex chromosomes. We also used the Genome in a Bottle (GIAB, https://www.nist.gov/programs-projects/genome-bottle) “difficult regions” track to check for parts of the genome where sequencing, mapping, and variant calling are highly challenging due to complex structures, repetitive elements, or alignment artifacts ^53^. We downloaded the ENCFF356LFX exclusion list (https://www.encodeproject.org/files/ENCFF356LFX/) and used the Bioconductor GenomicRanges package ^54^ to identify variants contained in the ENCODE blacklist^55^. We used the biomaRt package ^56^ to identify pseudo-genes on the sex chromosomes. We also checked the latest Genome Reference Consortium (GRC, https://www.ncbi.nlm.nih.gov/grc/human) patch for hg38 reference genome updates.

## Results

Among 6,156 WGS samples across eight data sets we constructed 2,071 case-parent trios (Supplementary Table 1). Quality control yielded numbers of variants to be tested in the respective cohorts between 6.8 and 9.8 million, with the exception of the African ancestry cohorts having more than 14 million variants (Supplementary Table 2) and reduced the number of trios included in the meta-analysis to 1,922 (Supplementary Table 3). The meta-analysis returned results for 18,261,298 distinct SNPs, including four SNPs in three distinct regions showing genome-wide significant differences in effects sizes between the sexes at a chosen genome-wide significance threshold of 1 × 10^−8^ (Figure 1, Table 1, Supplementary Figure 1).

**Figure 1:**
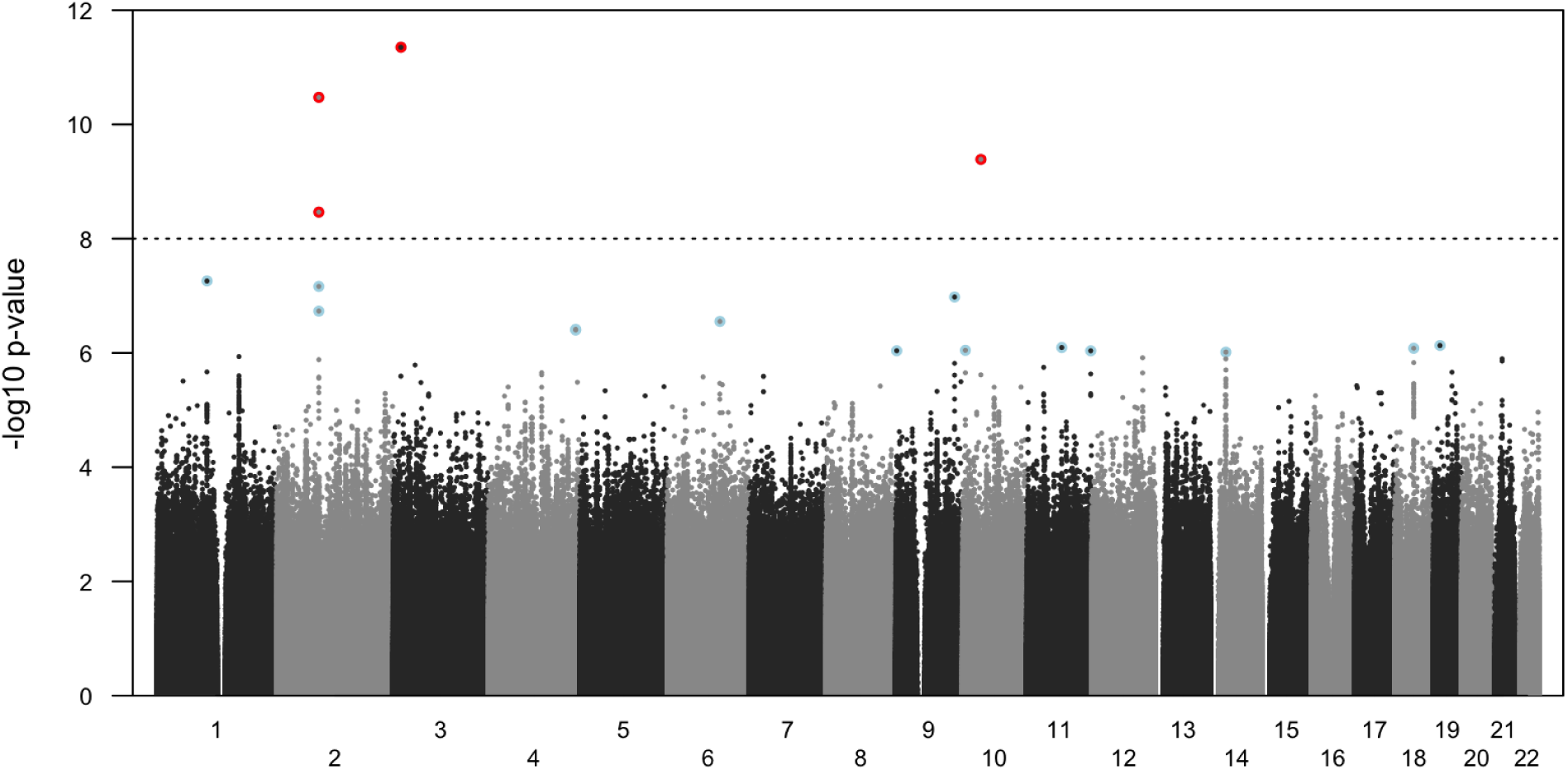
The Manhattan plot from the eight studies combined meta-analysis, testing the 1 degree of freedom SNP *×* Sex interaction in the conditional logistic gTDT model. The dashed horizontal line is located at p = 1 × 10^−8^, indicating genome-wide significance assuming 5 million independent SNPs. Highlighted SNPs are listed in Table 1. The four variants passing the genome-wide significance threshold are shown in red. Variants with a meta-analysis p-value less than 1 × 10^−6^ but not achieving genome-wide significance are shown in light blue.

The most significant SNP rs74520055 (p = 4.5*×*10^−12^ after genomic control correction with *λ* = 0.867, Supplementary Figure 2) at hg38 location 16,366,864 on chromosome 3 had a MAF of 11% in the first Latino cohort and was absent from all other OFC cohorts, despite having a non-zero MAF in all gnomAD populations. Male carriers had a substantially higher OFC risk than female carriers in this population, with a gTDT interaction parameter estimate of 4.04, corresponding to a relative risk ratio (RRr) of 56.7.

The second most significant SNP showing a difference in effects sizes between the sexes was rs2715199 at location 87,896,956 on chromosome 2 (p = 3.4*×*10^−11^, MAF = 6%). This marker was monomorphic in the two African ancestry populations but polymorphic in the six other cohorts with the same direction of effect (e.g., the parameter for the interaction in the gTDT conditional logistic regression model). In the European trios the effect was estimated to be -1.59 (p = 4.6 *×*10^−3^, MAF = 5%), in the Latino trios the effect was estimated to be -1.32 (p = 9.9 *×*10^−3^, MAF = 7%), in the Asian trios the effect was estimated to be -2.80 (p = 4.3 *×*10^−2^, MAF = 3%), in the first set of Filipino trios the effect was estimated to be -3.24 (p = 5.2 *×*10^−6^, MAF = 5%), in the second set of Filipino trios the effect was estimated to be -1.31 (p = 0.040, MAF = 4%) and in the second Latino cohort the effect was -1.15 (p = 0.016, MAF = 7%). In the meta-analysis the combined interaction parameter estimate was -1.54, corresponding to a RRr of 4.65 comparing female to male carriers. The SNP rs62140546 less than 33kb upstream of this finding also achieved genome-wide significance in the meta-analysis (p = 3.4*×*10^−9^, MAF = 6%), with an RRr of 5.42 comparing female to male carriers, which was very similar to rs2715199. In addition, two more markers in this region near 87.9Mb on chromosome 2 showed evidence for a sex-specific effects with roughly the same estimates of risk as the other markers in this region: rs199947305 (p = 6.8*×*10^−8^, MAF = 9%) and rs9647005 (p = 1.9*×*10^−7^, MAF = 8%).

Lastly, SNP rs3970232 on chromosome 10, observed in six studies, also achieved genome-wide signifi-cance in the meta-analysis (p = 4.1*×*10^−10^, MAF = 45%), albeit with a more moderate effect size in comparison to the previously described markers: the combined interaction parameter estimate was 0.64, corresponding to a relative risk ratio of 1.89 comparing male to female carriers. On the other hand, in comparison to these other variants, the minor allele frequency was much higher, increasing the power to detect more modest sex-specific effects.

However, additional QC specifically interrogating potential sequencing artifacts revealed concerns for the majority of variants listed in Table 1, including all four variants passing the genome-wide significance threshold of 1 × 10^−8^ (Supplementary Table 4). The most significant SNP rs74520055 on chromosome 3 (p = 4.5*×*10^−12^) was only observed in the Latino cohort with a MAF of 11%, despite having a non-zero MAF in all gnomAD populations (https://gnomad.broadinstitute.org) ^57,58^. The variant however failed the Allele-Specific Variant Quality Score Recalibration (AS VQSR) filter, a gnomAD QC metric used to distinguish true genetic variants from sequencing artifacts. Of note, all gnomAD populations also showed a much higher allele frequency among males than females. This presumably is a sequencing artifact due to the existence of an RFTN1 pseudo-gene on chromosome Y with more than 90% sequence identity to the corresponding chromosome 3 region and thus, Y-chromosome reads incorrectly map to the region with the marker rs74520055 (Supplementary Figure 3).

This was corroborated by several additional pieces of evidence. In our Latino cohort that generated this signal, the proportion of carriers of a variant allele among females is much lower than for males: 3.8% among daughters versus 40.4% among sons, and 6.9% among mothers versus 38.4% among fathers (Supplementary Figure 4) with parents generally not OFC affected. The SNP fails a test for Hardy-Weinberg equilibrium among male probands (p = 0.003) and fathers (p = 7 × 10^−5^) but not among female probands (p = 0.78) and mothers (p = 0.43). And while there are essentially no observed differences in relative read depths between carriers and non-carriers among female probands (fold change estimate 1.01, p = 0.83, 95% CI 0.85 – 1.20) and mothers (fold change estimate 1.01, p = 0.93, 95% CI 0.93 – 1.10) the observed relative read depth fold change at rs74520055 was 1.15 (p = 3.2 × 10^−6^, 95% CI 1.09 – 1.22) and 1.16 (p = 9.9 × 10^−11^, 95% CI 1.11 – 1.22) comparing carriers to non-carriers among male probands and fathers, respectively (Supplementary Figure 4). Lastly, 15 samples had a missing genotype in the VCF file due to low sequencing quality (GQ *<* 20), all were male (2 sons and 13 fathers).

The observed rs74520055 signal (RRr = 56.7) stems from the setting where a trio’s father is a heterozygote CT (HET) and the mother is a homozygous CC (HOM, Supplementary Table 5). Among 52 such trios with a male proband, 48 sons were HET. Among 33 such trios with a female proband, 32 daughters were HOM. It thus appears that most of the HETs are incorrect sequencing calls. There are very few HETs among the mothers and daughters indicating a low true MAF in the population, estimated at 3.0% from females only. If there is a variant on a father’s Y chromosome leading to a HET call for rs74520055, then this Y variant will be inherited if the proband is male, i.e. the HET will be present in the father and the son. If the proband is female, the father’s X chromosome will be passed to the daughter, and thus, the HET will only be present in the father (Supplementary Figure 3). The four male HOM probands and the one HET female proband among the trios with a HET father and a HOM mother were likely true Mendelian events.

The second most significant SNP rs2715199 on chromosome 2 (p = 3.4*×*10^−11^) was polymorphic in six cohorts with the same direction of effect. It also showed appreciable variation in all gnomAD populations with a substantially higher allele frequency among females than males. This presumably is another sequencing artifact as there exists a region on the X chromosome with more than 99% sequence identity to the chromosome 2 region and thus, X-chromosome reads incorrectly aligning to the corresponding chromosome 2 region with the marker rs2715199 (Supplementary Figure 5).

The observed rs2715199 signal (RRr = 0.22, i.e. female to male RRr = 4.65) stems from the setting where a trio’s father is a HET and the mother is a HOM (Supplementary Table 6). For example, among 17 such trios with a male proband in the European cohort, all sons were HOM. Among 7 such trios with a female proband, all daughters were HET. The autosomal marker might actually be monomorphic in truth and the variant only present on the X chromosome. Then in this setting, all father’s rs2715199 HET calls are false. However, since a male proband always inherits the Y chromosome, the variant carrying X chromosome does not get passed on and therefore each son is a HOM. On the other hand, a female proband always inherits the variant carrying X chromosome from the father and therefore each daughter is a HET (Supplementary Figure 5).

Since each female has two X and each male only one X chromosome, one would expect more HETs among females in this setting. This is indeed the case: 13.0% HETs among daughters versus 5.3% among sons, and 11.2% among mothers versus 7.6% among fathers. Comparing rs2715199 carriers to non-carriers, we observed a 1.09 fold change (p = 0.002, 95% CI 1.03 – 1.15) in relative read depths across all four groups, and in addition, a relative read depths 1.23 fold change (p *<* 10^−10^, 95% CI 1.19 – 1.27) jointly comparing mothers and daughters to fathers and sons (Supplementary Figure 6). The other genome-wide significant SNP in this region of chromosome 2, rs62140546, exhibits similar statistics (Supplementary Figure 7 and Supplementary Table 7).

Lastly, SNP rs3970232 on chromosome 10 achieved genome-wide significance (p = 4.1*×*10^−10^). Its genomic locus is in the pericentromeric region of chromosome 10 (Supplementary Figure 1), less than 1Mb from the beginning of the centromere. Pericentromeric regions are enriched for highly homologous segmental duplications and repetitive sequences, making read mapping particularly challenging ^59^. Long-range LD is commonly observed in such regions due to reduced crossover recombination, which produces extended correlations between variants across large physical distances (e.g., Supplementary Figure 1), and consequently reduces fine-mapping resolution ^60–62^. SNP rs3970232 is included in the ENCODE blacklist ^55^ and failed several of our follow-up QC indicators (Supplementary Table 4).

## Discussion

Few papers in the literature have incorporated SNP *×* Sex interactions in studies of OFCs. Carlson et. al. ^22^ assessed SNP *×* Sex interactions in a population based study using 2,142 CL/P cases and 1,700 controls of European ancestry. Among loci with suggestive results (p *<* 10^−5^) in a joint 2df test of main effect and interaction, one SNP (rs72804706 on 10q21) achieved genome-wide significance for the SNP*×*Sex interaction and 16 more SNPs reached suggestive levels for the interaction (Table 2 in Carlson et. al. ^22^). Awotoye et. al. ^23^ conducted a GWAS of SNP *×* Sex interactions in a sub-Saharan African cohort, comprised of 1,019 OFC cases and 2,159 controls recruited from Ethiopia, Ghana, and Nigeria. Similarly, this study considered the SNP *×* Sex interaction results among the loci with suggestive results (p *<* 10^−5^) in the joint 2df test of both main effect and interaction, with one of them (rs2720555 on 8p22) reaching genome-wide significance for the SNP *×* Sex interaction (Appendix Table 2, Awotoye et. al. ^23^). However, none of these SNPs were validated in our meta-analyses (data not shown). In contrast to our multi-ethnic meta-analysis, these studies were only of European and African ancestry participants respectively, and thus the lack of validation may not be too surprising. These studies also did not conduct a post-analysis quality control, so it is unclear which of these reported variants might have been due to similar sequencing artifacts as we have observed in our study.

It is noteworthy that the vast majority of SNP *×* Sex interaction papers appear to focus on statistical methodology and biological interpretation^63–66^ while very few systematically investigate whether reported loci reside in low-mappability or blacklisted regions. Given the majority of our findings with an interaction p-value less than 10^−6^ had at least one potential concern and some of those, including all four variants passing the genome-wide significance threshold, appeared to be clear sequencing errors (Supplementary Table 4) highlights what we believe is a critical but commonly omitted step in the analysis of sex-specific genetic effects. That said, several of our findings with an interaction p-value less than 10^−6^ did not reveal apparent artifacts and showed biologically plausible effect sizes, consistent with the expected polygenic architecture of OFCs. Several of these suggestive variants act as eQTLs for genes critically relevant to craniofacial development, most notably rs6568608 on chromosome 6 for *FIG4* and *MICAL1*, which are associated with syndromes featuring skeletal abnormalities and CL/P^67,68^ and rs3793516 on chromosome 9 for *SMARCA2* linked to syndromes featuring high palate and other craniofacial features^69^. These leads are further supported by predicted impacts on transcriptional factor binding for key developmental regulators like *HOXB7*, *OTX2* and *IKZF1* ^70^ marking them as high-priority candidates for future validation of sex-specific risk.

A critical challenge in large-scale meta-analyses of WGS data stems from heterogeneity in sequencing, variant-calling and QC pipelines, which can introduce batch effects that can generate spurious association results if not properly addressed. Harmonizing data processing across studies has been shown to reduce technical variability and improved genotype concordance rates have been reported when best-practice QC filters are applied across datasets ^71^. Analyses of chromosome X in WGS meta-analyses are even more vulnerable to batch effects, owing to fundamental biological and technical features distinguishing X from the autosomes. Sex-specific ploidy differences, variable treatment of pseudoautosomal regions, and reduced effective coverage in males amplify the impact of differences in pipelines across cohorts. Large benchmarking efforts and consortia have consistently shown that pipeline heterogeneity explains a disproportionate fraction of technical variance on chromosome X, leading to sex-biased missingness, allele frequency distortion, and reduced variant concordance compared with autosomes ^71^. Analyses from population-scale resources such as gnomAD further demonstrate that X-chromosome variant quality and callability are markedly improved when genomes are processed through a unified, ploidy-aware pipeline with harmonized QC filters, whereas heterogeneous processing introduces systematic artifacts that can mimic or obscure true X-linked signals ^72^. Recent UK Biobank studies similarly report substantial sex-dependent genotype missingness and allele frequency discrepancies on chromosome X, underscoring the sensitivity of X-linked analyses to technical batch effects in large cohorts ^73^. Fortunately, sex-aware quality control procedures to identify, understand and correct technical artifacts in genotype calls for the sex chromosomes in next-generation sequencing data is being addressed in an active research area ^74,75^. However, in addition to the higher potential of technical artifacts across cohorts, the analysis framework for chromosome X variants also needs to be altered to account for sex-specific ploidy and inheritance patterns. We have thus decided to limit our analyses to autosomal variants in this manuscript and will address chromosome X variants separately in a future study.

## Data Availability

Participant sequencing and basic phenotype data are available from the database of Genotypes and Phenotypes (dbGaP) under the accesion numbers phs001168.v1.p1 (study 1), phs001420.v1.p1 and phs001420.v1.p1 (2), phs001997.v1.p1 (3 and 4), phs002595.v1.p1 (5) and phs001997.V1.p1 (6). Studies 7 and 8 are currently under submission to dbGaP (accession number to be determined). Additional phenotype information is available from the FaceBase website for studies 1, 2, 5, 7, and 8 (https://doi.org/10.25550/5A-FJBJ and https://doi.org/10.25550/56-ES6P).

## Acknowledgements

Samples from studies 1, 2, 5, 7 and 8 are participants drawn from the multi-ancestry Pittsburgh Orofacial Cleft study (POFC) with funding for recruitment supported by R01 DE016148 and R01 DE008559. Study participants were recruited per the collaborative protocol established by the POFC coordinating center, with the approval of the University of Pittsburgh Biomedical IRB (FWA00006790), as well as the approval of site-specific ethics committees. Samples from study 4 were drawn from the International Consortium to Identify Genes and Interactions Controlling Oral Cleft supported by U01 DE018993. Each participating institution reviewed and approved research protocols for recruiting human subjects, and US institutions reviewed and approved protocols of their foreign collaborators. After this International Cleft Consortium was formed in 2007, each individual IRB reviewed and approved monitored data sharing protocols administered through dbGaP (JHU FWA 00000287). Following the establishment of the African Craniofacial Anomalies Research Network in 2012, IRB approval was obtained from the University of Lagos in Nigeria, Addis Ababa University in Ethiopia and Kwame Nkurumah University of Science and Technology in Ghana. Sequencing of samples was supported by grants X01 HL132363 (study 1), X01 HL136465 and X01 DE030062 (2), X01 HL140516 (3 and 4), X01 HD100701 (5), X01-HL140516-01 (6) and X01 DE032472 (7 and 8). KK, SB, NM, DR, THB, MLM and IR were supported by R01 DE031855. MM and ZE were also supported by R01 DE032319. ZE was further supported by T90 DE030853. AB was supported by R01 DE028300. This research was supported in part by the Intramural Research Program of the National Institutes of Health (NIH). The contributions of the NIH authors are considered Works of the United States Government. The findings and conclusions presented in this paper are those of the authors and do not necessarily reflect the views of the NIH or the U.S. Department of Health and Human Services.

## Supplementary Materials

**Supplementary Table 1:** The number of samples, case-parent trios, probands and parents included in the multi-VCFs for eight different datasets we included in the quality control pipelines and analyses. In dataset 6 two cases were siblings yielding two separate trios. In datasets 7 and 8 some trios were extracted from extended families. All other datasets contain complete trios with each member being sequenced once.

|  | Ethnicity | Samples | Trios | Probands | Parents |
| --- | --- | --- | --- | --- | --- |
| 1 | European | 1,113 | 371 | 371 | 742 |
| 2 | Latino | 795 | 265 | 265 | 530 |
| 3 | African | 411 | 137 | 137 | 274 |
| 4 | Asian | 375 | 125 | 125 | 250 |
| 5 | Filipino | 1,110 | 370 | 370 | 740 |
| 6 | African | 452 | 152 | 152 | 300 |
| 7 | Filipino | 859 | 302 | 302 | 524 |
| 8 | Latino | 1,041 | 349 | 349 | 692 |
|  | Total | 6,156 | 2,071 | 2,071 | 4,052 |

**Supplementary Table 2:** The number of variants in the released VCF files (Pre QC) and number of variants left in the VCFs after using VCFtools to remove multi-allelic sites and indels, filter sites with labels other than ‘PASS’, and removing variants with low mean depth values or low quality values (Basic QC). The number of SNPs not passing the filters for missingness (Missing), Hardy-Weinberg equilibrium (HWE) and minor allele frequency (MAF) are also given (a SNP can fail multiple filters). The final number of variants used in the analyses is given in the final column (Total). Sequencing and VCF generation was carried out at the Broad Institute except for dataset 6 which was conducted at the Center for Inherited Disease Research, using a different VCF release protocol. Datasets 7 and 8 were generated from a larger dataset of subjects with mixed ethnicities and procesed on CAVATICA in the cloud. The VCF available had already been pre-screened for variants with less than 1% MAF.

|  | Ethnicity | Pre QC | Basic QC | Missing | HWE | MAF | Total |
| --- | --- | --- | --- | --- | --- | --- | --- |
| 1 | European | 53,404,248 | 35,269,929 | 72,634 | 4,349 | 26,961,827 | 8,201,682 |
| 2 | Latino | 44,586,948 | 29,181,084 | 37,763 | 8,350 | 19,333,771 | 9,791,103 |
| 3 | African | 42,679,800 | 27,294,048 | 50,993 | 2,116 | 13,068,663 | 14,319,481 |
| 4 | Asian | 26,555,387 | 16,788,744 | 29,669 | 1,965 | 9,241,079 | 7,354,165 |
| 5 | Filipino | 35,827,309 | 24,467,046 | 311,984 | 29,438 | 16,263,011 | 7,866,804 |
| 6 | African | 58,188,374 | 49,868,598 | 5,883,543 | 21,605 | 29,433,009 | 14,250,368 |
| 7 | Filipino | 14,557,705 | 9,153,901 | 146,057 | 18,654 | 2,032,716 | 6,767,554 |
| 8 | Latino | 14,746,625 | 8,967,521 | 145,291 | 46,044 | 1,015,843 | 7,760,343 |

**Supplementary Table 3:** The number of case-parent trios, and the number of male/female probands in the trios, passing all QC filters and included in sex-specific gTDT, for the eight different data sets.

|  | Ethnicity | Trios | Male | Female |
| --- | --- | --- | --- | --- |
| 1 | European | 341 | 218 | 123 |
| 2 | Latino | 259 | 149 | 110 |
| 3 | African | 129 | 65 | 64 |
| 4 | Asian | 124 | 84 | 40 |
| 5 | Filipino | 354 | 234 | 120 |
| 6 | African | 126 | 69 | 57 |
| 7 | Filipino | 287 | 167 | 120 |
| 8 | Latino | 302 | 180 | 122 |
|  | Total | 1,922 | 1,166 | 756 |

**Supplementary Table 4:** Variants with the 1 degree of freedom SNP*×*Sex interaction p-values listed in Table 1. BLAT sequence similarity to other chromosomes (B, shown as percentage/chromosome), self-chain results (S, chromosome shown), ENCODE blacklist inclusion (E), known pseudo-genes (P) or other reasons (O) indicate questionable results. SNP rs73869151 is in and SNPs rs9654290 and rs60598949 are next to “difficult regions” defined by the Genome in a Bottle Consortium. SNP rs3970232 is in a repetitive region near the centromere and self-chains to multiple chromosomes including chromosome Y. SNP rs184424374 is in a Genome Reference Consortium sequence patch.

| SNP | CHR | Position | P-value | B | S | E | P | O |
| --- | --- | --- | --- | --- | --- | --- | --- | --- |
| rs11488076 | 1 | 104,390,587 | 5.5e-08 | 85/X |  |  |  |  |
| rs199947305 | 2 | 87,844,559 | 6.8e-08 | 99/X | X |  |  |  |
| rs62140546 | 2 | 87,864,648 | 3.4e-09 | 99/X | X |  |  |  |
| rs9647005 | 2 | 87,877,885 | 1.9e-07 | 99/X | X |  |  |  |
| rs2715199 | 2 | 87,896,956 | 3.4e-11 | 99/X | X |  |  |  |
| rs74520055 | 3 | 16,366,864 | 4.5e-12 | 90/Y | Y |  | + |  |
| rs73869151 | 4 | 181,496,591 | 4.0e-07 |  |  |  |  | + |
| rs9654290 | 4 | 181,496,688 | 3.9e-07 |  |  |  |  | + |
| rs6568608 | 6 | 109,778,879 | 2.8e-07 |  |  |  |  |  |
| rs3793516 | 9 | 2,174,095 | 9.1e-07 |  |  |  |  |  |
| rs60598949 | 9 | 122,366,992 | 1.0e-07 |  |  |  |  | + |
| rs57238180 | 10 | 6,606,368 | 9.0e-07 |  |  |  |  |  |
| rs3970232 | 10 | 38,788,107 | 4.1e-10 |  | Y/+ | + |  | + |
| rs56927892 | 11 | 73,155,016 | 8.0e-07 |  |  |  |  |  |
| rs35838389 | 11 | 133,403,398 | 9.2e-07 |  |  |  |  |  |
| rs4981892 | 14 | 31,724,818 | 9.7e-07 |  |  |  |  |  |
| rs9945692 | 18 | 39,903,998 | 8.3e-07 |  |  |  |  |  |
| rs184424374 | 19 | 142,49,017 | 7.4e-07 |  | 17 |  |  | + |

**Supplementary Table 5:** The number of observed chromosome 3 rs74520055 trio genotype combinations (father/mother/child) observed in the Latino cohort for trios with a male (# M) or with a female (# F) proband, and the respective frequencies (% M, % F). The genotype combinations driving the signal (RRr = 56.7 comparing male to female relative risks) are highlighted with a grey background.

|  | # M | # F | % M | % F |
| --- | --- | --- | --- | --- |
| CC/CC/CC | 77 | 62 | 54.6 | 59.6 |
| CC/CC/CT | 3 | 0 | 2.1 | 0.0 |
| CC/CT/CC | 3 | 4 | 2.1 | 3.8 |
| CC/CT/CT | 1 | 1 | 0.7 | 1.0 |
| CT/CC/CC | 4 | 32 | 2.8 | 30.8 |
| CT/CC/CT | 48 | 1 | 34.0 | 1.0 |
| CT/CT/CC | 0 | 2 | 0.0 | 1.9 |
| CT/CT/CT | 3 | 2 | 2.1 | 1.9 |
| CT/CT/TT | 1 | 0 | 0.7 | 0.0 |
| TT/CC/CT | 1 | 0 | 0.7 | 0.0 |

**Supplementary Table 6:** The number of observed chromosome 2 rs2715199 trio genotype combinations (father/mother/child) observed in the European cohort for trios with a male (# M) or with a female (# F) proband, and the respective frequencies (% M, % F). The genotype combinations driving the signal (RRr = 4.65 comparing female to male relative risks) are highlighted with a grey background.

|  | # M | # F | % M | % F |
| --- | --- | --- | --- | --- |
| AA/AA/AA | 171 | 99 | 82.2 | 80.5 |
| AA/AG/AA | 8 | 8 | 3.8 | 6.5 |
| AA/AG/AG | 11 | 9 | 5.3 | 7.3 |
| AG/AA/AA | 17 | 0 | 8.2 | 0 |
| AG/AA/AG | 0 | 7 | 0.0 | 5.7 |
| AG/AG/AA | 1 | 0 | 0.5 | 0.0 |

**Supplementary Table 7:** The number of observed chromosome 2 rs62140546 trio genotype combinations (father/mother/child) observed in the European cohort for trios with a male (# M) or with a female (# F) proband, and the respective frequencies (% M, % F). The genotype combinations driving the signal (RRr = 5.42 comparing female to male relative risks) are highlighted with a grey background.

|  | # M | # F | % M | % F |
| --- | --- | --- | --- | --- |
| AA/AA/AA | 149 | 80 | 70.0 | 68.4 |
| AA/AT/AA | 11 | 8 | 5.2 | 6.8 |
| AA/AT/AT | 24 | 16 | 11.3 | 13.7 |
| AT/AA/AA | 27 | 0 | 12.7 | 0.0 |
| AT/AA/AT | 0 | 11 | 0.0 | 9.4 |
| AT/AT/AA | 2 | 0 | 0.9 | 0.0 |
| AT/AT/AT | 0 | 2 | 0.0 | 1.7 |

**Supplementary Figure 1:**
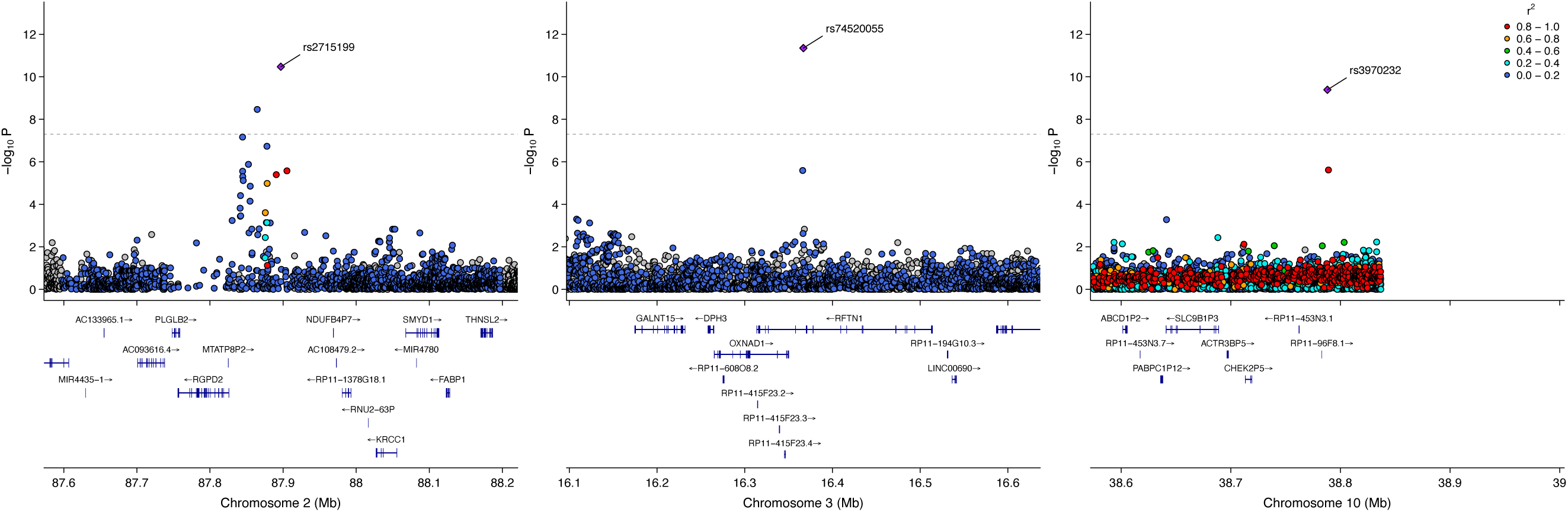
Locuszoom plots for the chromosome 2 (**left**), chromosome 3 (**middle**) and chromosome 10 (**right**) loci. The long-range LD pattern near the chromosome 10 locus is commonly observed near centromeres due to reduced crossover recombination, producing extended correlations between variants across large physical distances.

**Supplementary Figure 2:**
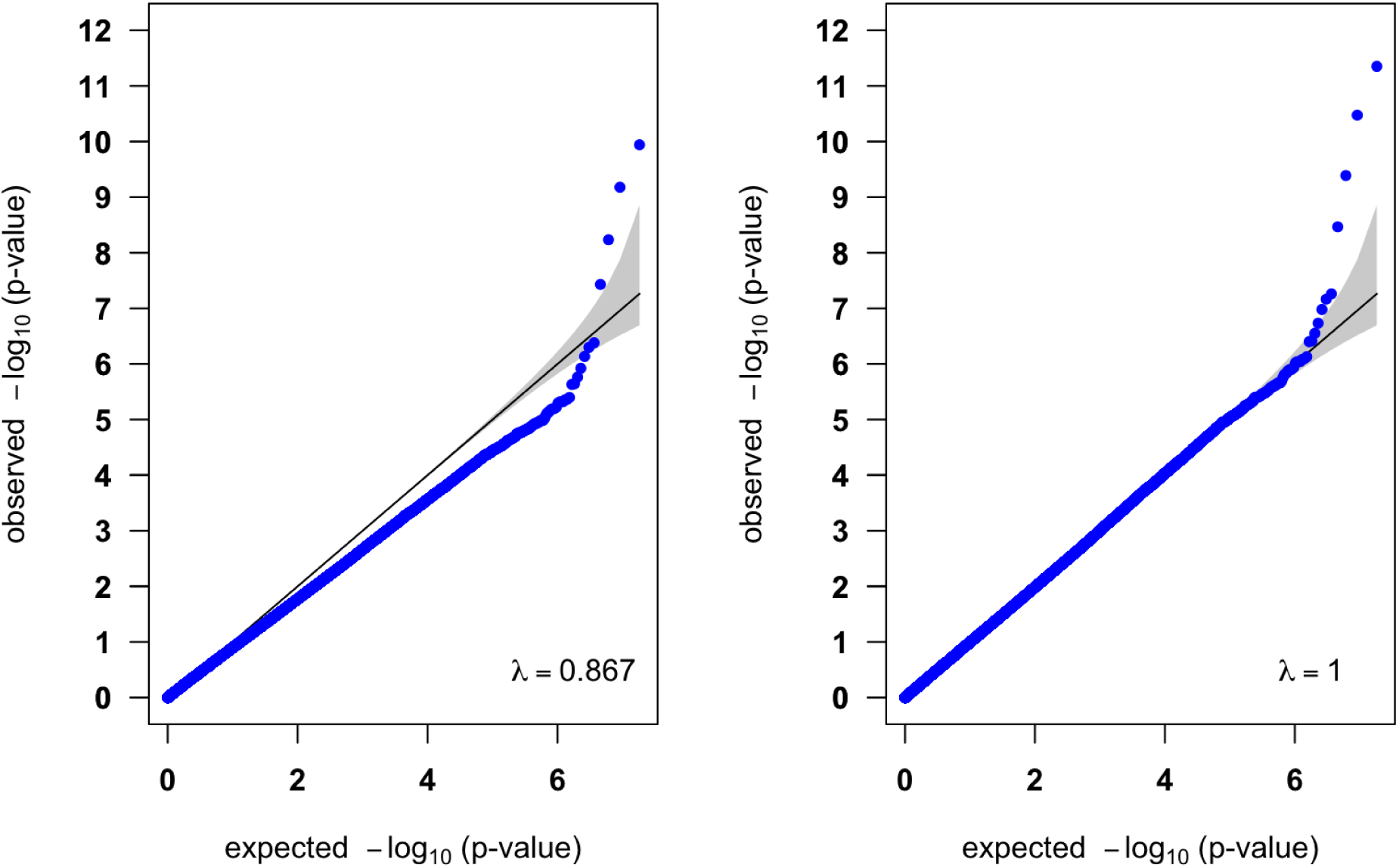
QQ-plots for the 18,261,298 p-values from the eight studies meta-analysis before (**left**) and after (**right**) genomic control correction.

**Supplementary Figure 3:**
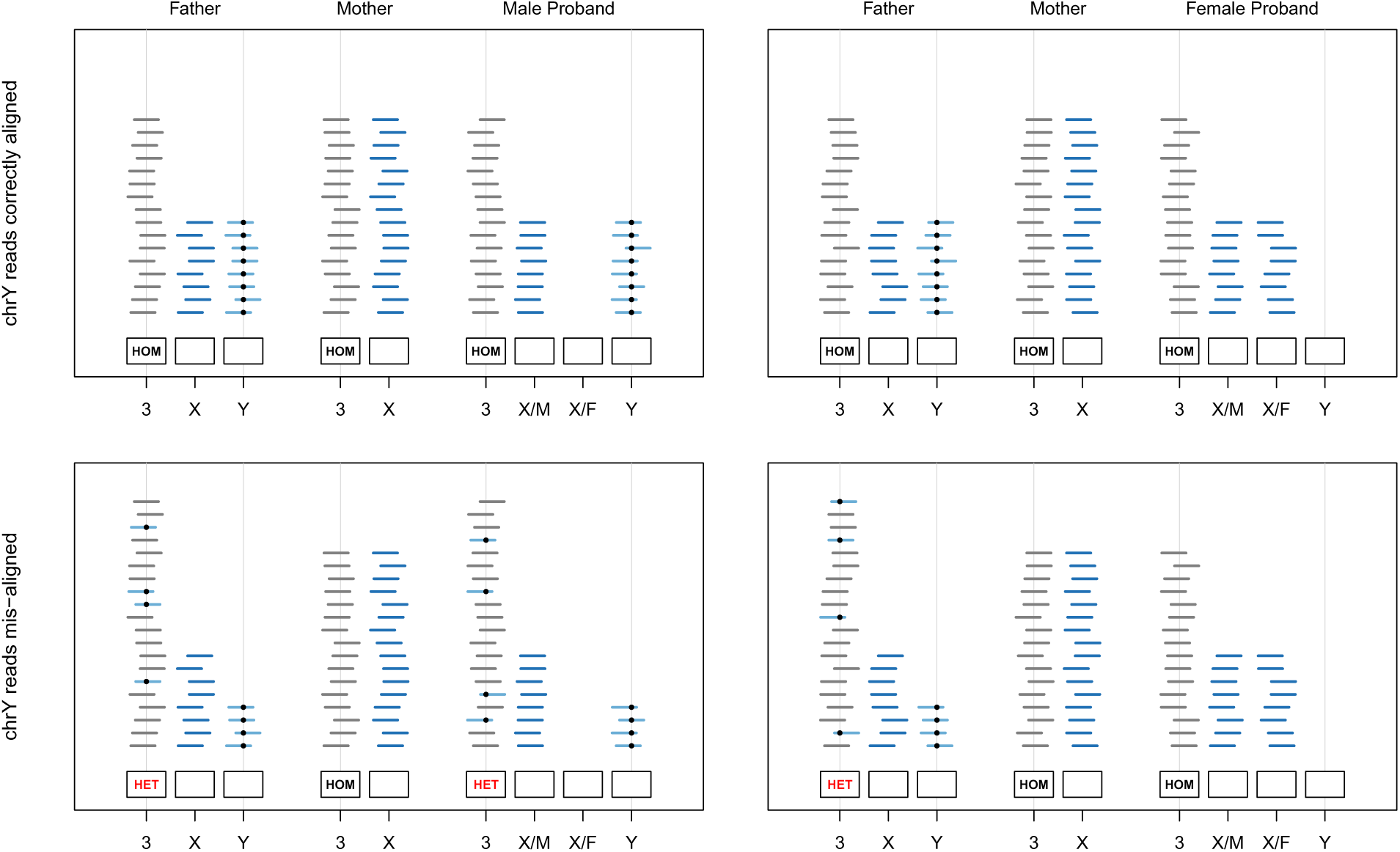
Hypothesized read mis-alignment leading to the chromosome 3 artifact. Chomosome 3 reads are shown in grey, X chromosome reads are shown in dark blue, Y chromosome reads are shown in light blue. X/M indicates an X chromosome inherited from the mother, X/F indicates an X chromosome inherited from the father. The hg38 chromosome 3 position 16,366,864 and the corresponding location in the pseudogene on the Y chromosome are indicated by vertical lines. Due to sequence similarity, chromosome Y reads can be mapped to chromosome 3 for fathers and male probands. If all trio members are REF homozygous (HOM) but a variant (black dot) is present on the Y chromosome (top row), chromosome Y read misalignment can lead to heterozygous (HET) genotype calls in fathers (bottom row) and male probands (bottom left), but not female probands (bottom right). Thus, it appears that rs74520055 variants are preferentially transmitted to male probands, leading to a significant SNP *×* Sex interaction.

**Supplementary Figure 4:**
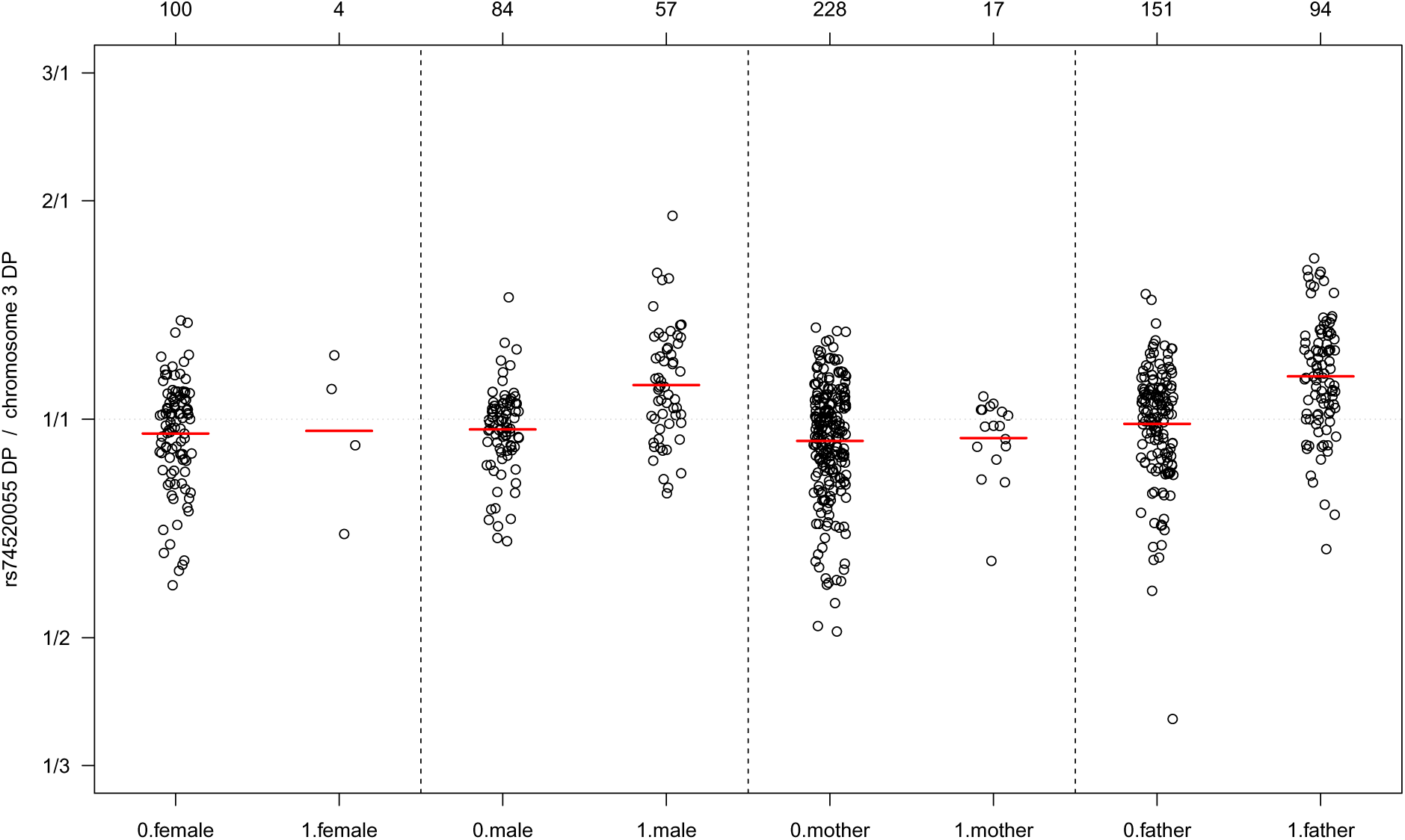
The sample read depths observed at SNP rs74520055 relative to the respective sample average chromosome 3 read depths between carriers of a variant allele (1) and homozygous reference allele carriers (0) for female probands, male probands, mothers and fathers, with geometric means shown in red. The number of samples in each group is shown at the top of the plot. One male proband and one father were reported as homozygous carriers of the variant allele, all other carriers were reported heterozygotes.

**Supplementary Figure 5:**
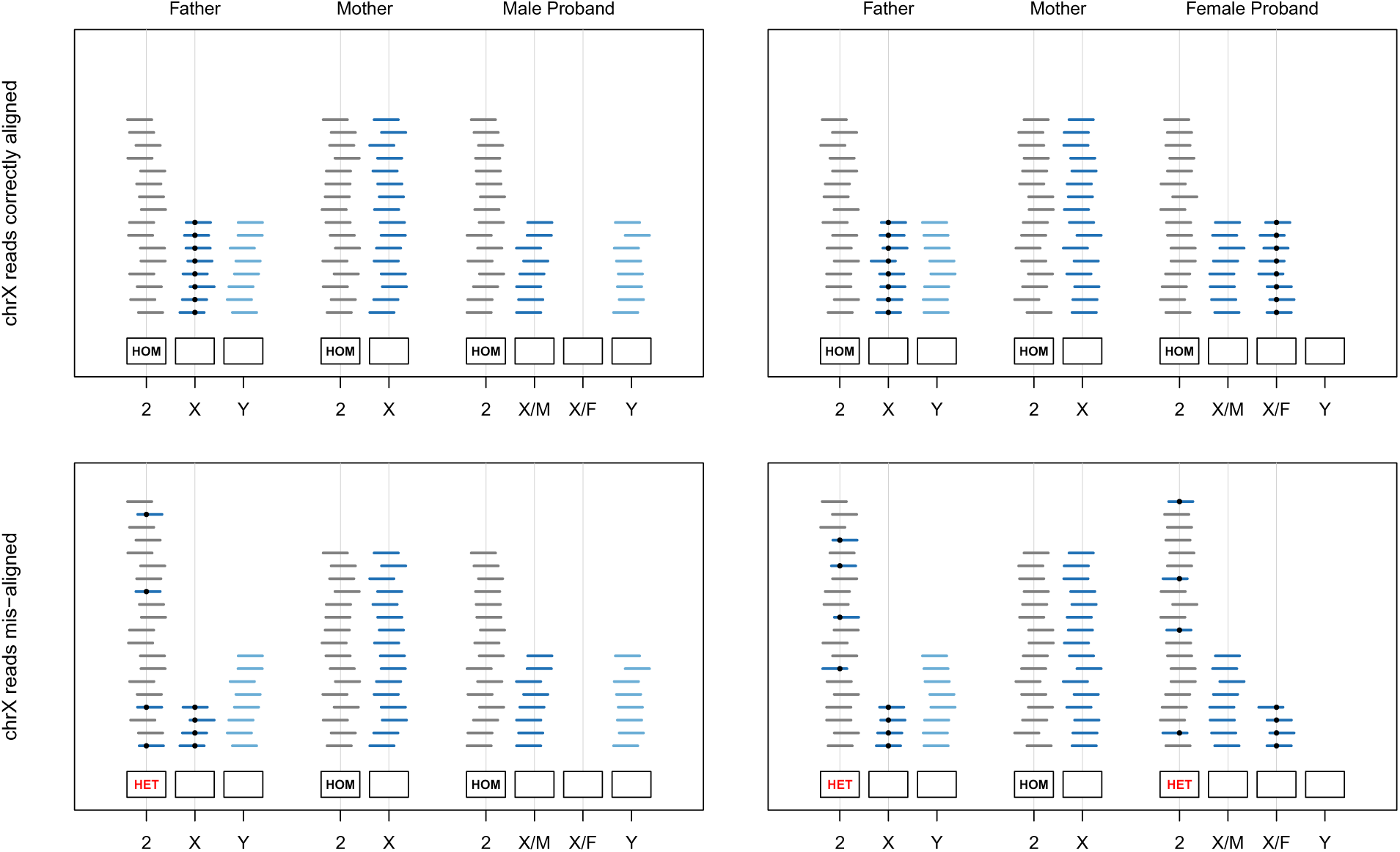
Hypothesized read mis-alignment leading to the chromosome 2 artifact. Chomosome 2 reads are shown in grey, X chromosome reads are shown in dark blue, Y chromosome reads are shown in light blue. X/M indicates an X chromosome inherited from the mother, X/F indicates an X chromosome inherited from the father. The hg38 chromosome 2 position 87,896,956 and the corresponding location in the nearly identical region on the X chromosome are indicated by vertical lines. Due to sequence similarity, chromosome X reads can be mapped to chromosome 2 for all trio members. If all trio members are REF homozygous (HOM) but a variant (black dot) is present on the X chromosome in a father, also inherited by a female proband (top row), chromosome X read misalignment can lead to heterozygous (HET) genotype calls in fathers (bottom row) and female probands (bottom right), but not male probands (bottom left). Thus, it appears that rs2715199 variants are preferentially transmitted to female probands, leading to a significant SNP *×* Sex interaction.

**Supplementary Figure 6:**
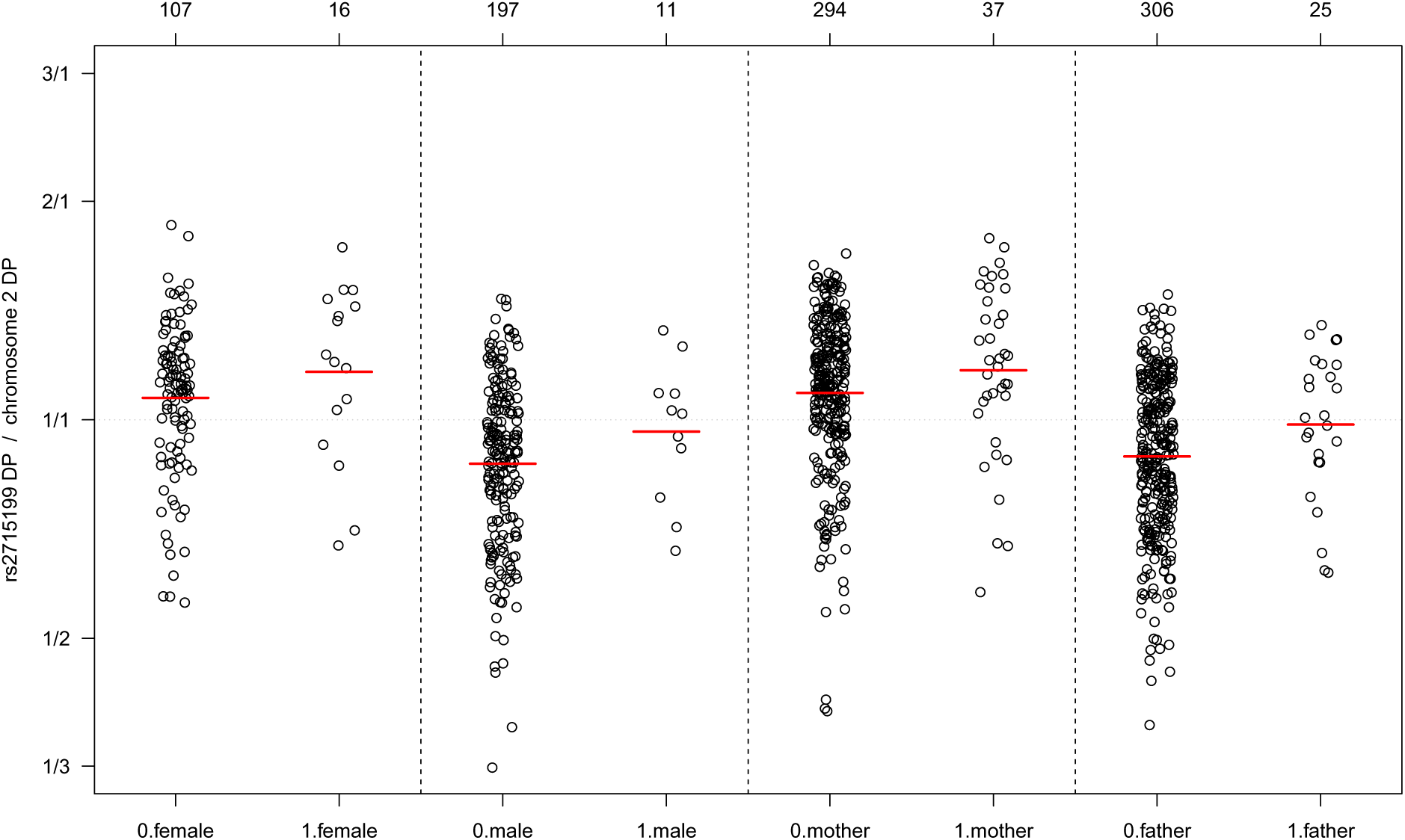
The sample read depths observed at SNP rs2715199 relative to the respective sample average chromosome 2 read depths between carriers of a variant allele (1) and homozygous reference allele carriers (0) for female probands, male probands, mothers and fathers, with geometric means shown in red. The number of samples in each group is shown at the top of the plot.

**Supplementary Figure 7:**
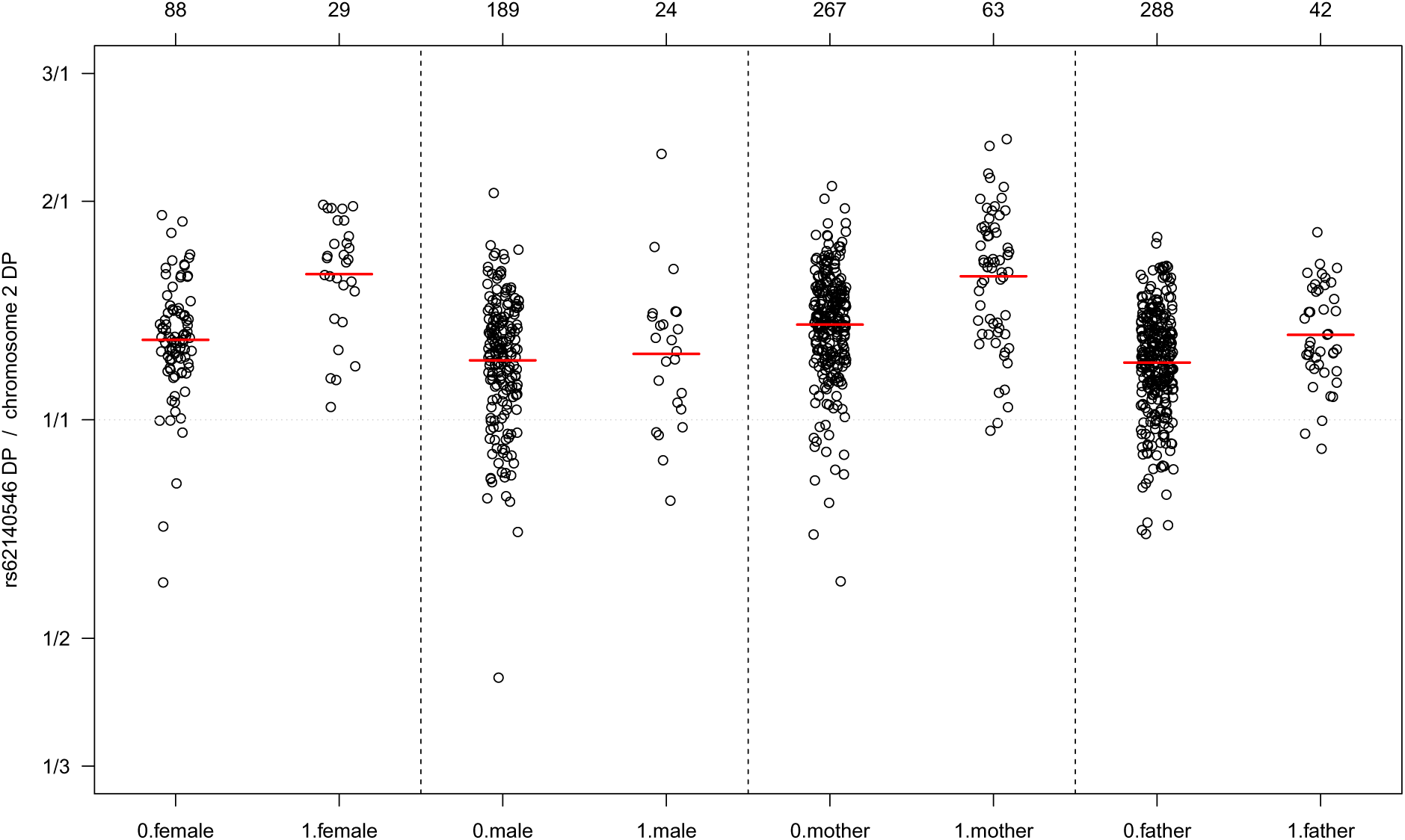
The sample read depths observed at SNP rs62140546 relative to the respective sample average chromosome 2 read depths between carriers of a variant allele (1) and homozygous reference allele carriers (0) for female probands, male probands, mothers and fathers, with geometric means shown in red. The number of samples in each group is shown at the top of the plot.

